# Remote Perioperative Symptom Monitoring via Smartphone is Feasible: Evidence from the Personalized Prediction of Persistent Postsurgical Pain (P5) study of 2,500 surgical patients

**DOI:** 10.1101/2025.07.27.25332242

**Authors:** Madelyn R. Frumkin, Gabrielle R. Messner, Katherine J. Holzer, Ziqi Xu, Thomas L. Rodebaugh, Haley Bernstein, Karen Frey, Saivee Ahuja, Jorin Hanson, Chenyang Lu, Simon Haroutounian

## Abstract

Ecological momentary assessment (EMA) holds promise for perioperative monitoring, yet large-scale feasibility data are lacking. The Personalized Prediction of Persistent Postsurgical Pain (P5) study enrolled 2,500 adults undergoing major surgery at a single center. EMA consisting of 15 items was administered three times daily via smartphone. Participants were not directly incentivized for EMA compliance nor excluded for non-compliance. Approximately 90% of participants completed at least some EMA. Average preoperative compliance was 66% (Median=79%) and average postoperative compliance was 60% (Median=71%) in the first 30 days after surgery. Postoperative compliance differed by surgical site, with lowest compliance among vascular and cardiothoracic patients. Demographic characteristics, including race, insurance status, and education, were associated with compliance. Overall, perioperative EMA appears feasible. Appropriate handling of missing data is critical to ensure models are generalizable to individuals who hold marginalized identities.

---

Ecological Momentary Assessment (EMA) facilitates frequent capture of patient-reported symptoms and experiences via smartphone^1^. Repeated measurements within daily life reduce recall bias associated with traditional patient-reported outcome measures and improve ecological validity^2^. Limited research suggests EMA may be useful for monitoring postoperative recovery and identifying opportunities for early intervention^3–6^. Preoperative EMA also facilitates the development of novel features (e.g., symptom trajectories and dynamics) that may improve prediction of surgical outcomes, including persistent post-surgical pain^4, 5^. However, adoption of remote perioperative monitoring via smartphone is hindered by a lack of largescale studies supporting feasibility.

The Personalized Prediction of Persistent Postsurgical Pain (P5) study is a single-center, prospective study that aims to examine whether multimodal data, including EMA, improve prediction of persistent postsurgical pain (see ^7^). In the current planned secondary analysis, we aimed to examine compliance with EMA in this largescale surgical cohort.

P5 included adults aged 18 to 75 undergoing major surgery (surgery duration >1h with a planned overnight hospital stay). Access to a smartphone or tablet was required for study participation, although individuals who had access to someone else’s device (e.g., spouse) could be included. Individuals unable to speak or read English were excluded. The study was approved by the university’s Institutional Review Board. Participants were compensated $50 for completion of the preoperative visit and baseline surveys. Participants were not compensated based on EMA compliance. EMA included 15 items administered 3 times per day. Detailed methods are provided in the Supplementary Materials.

A total of 2500 participants signed consent, and 107 participants (4.3%) became ineligible prior to surgery for reasons including canceled or changed surgery and non-completion of baseline measures. Another 64 participants (2.6%) asked to withdraw prior to surgery. Of the remaining 2329 participants, 67% (n=1555) were female. Participants primarily identified their race as White/Caucasian (n=1752, 75.2%) or Black/African heritage (n=427, 18.3%) and were 53 years old on average (SD=13.8, Min=18, Max=75). Additional demographic and clinical characteristics of the sample are available in **Supplementary Table 2**.

Most participants (n=2100, 90%) completed some pre- or postoperative EMA. In univariate analyses (**Supplementary Table 2)**, participants who did not complete any EMA were older (Median age = 58.6 vs. 54.2) and more likely to have governmentally subsidized insurance (i.e., Medicaid). There were no other demographic or clinical differences between those who did and did not complete EMA. Average EMA response time was 89 seconds (Median = 65 seconds).

Average preoperative compliance was 66% (SD=33%, Median=79%). As shown in **Fig 1A**, the distribution of preoperative compliance was left-skewed (skewness=−0.88), suggesting high compliance was more common than low compliance. Preoperative EMA compliance did not differ by surgical site (**Fig 1B**).

**Fig 1.**
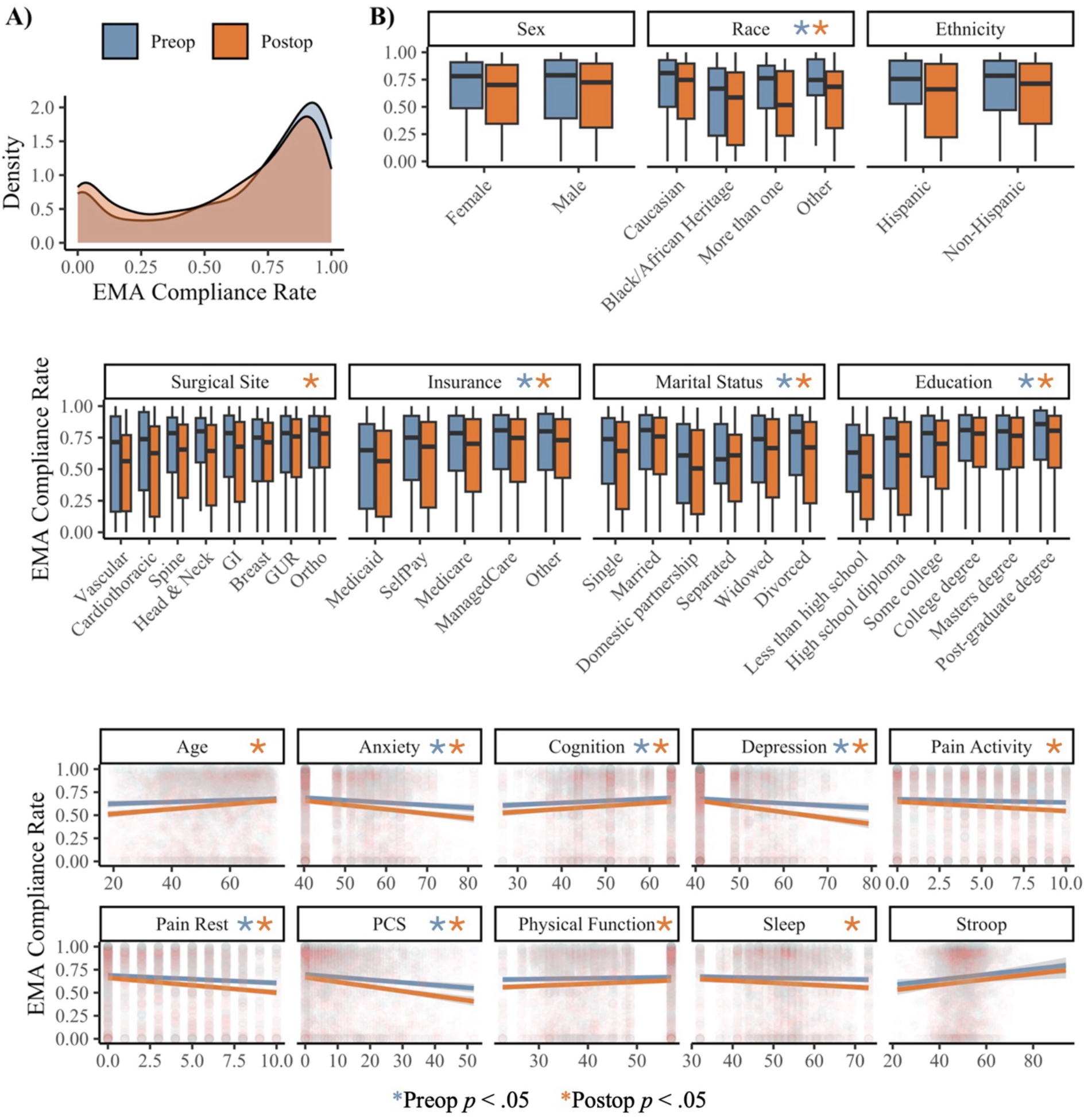
Pre- and post-surgery EMA compliance. **Panel A** shows compliance distributions across the full sample, including all participants who received at least 1 EMA. **Panel B** shows demographic and clinical correlates of EMA compliance, based on univariate analyses. Point estimates, significance, and effect sizes are available in the Supplementary Materials. PCS = Pain Catastrophizing Scale; GI = gastrointestinal; GUR = Genitourinary. In Panel B, anxiety, subjective cognition, depression, physical function, and subjective sleep impairment were assessed via PROMIS self-report measures; pain with activity and pain at rest were assessed via numeric rating scales.

Average postoperative compliance was 60% (SD=33%, Median=71%, skewness=−0.62). Postoperative EMA compliance did differ by surgical site (η^2^=0.029, p < .001), with below average postoperative compliance observed among patients who underwent vascular and cardiothoracic procedures (*M*=49-51%, Median=56-63%).

Several clinical characteristics were univariately associated with EMA compliance, although effect sizes were small (|r| ≤ .18; Figure S1-S2). Preoperative pain catastrophizing was the strongest correlate of both preoperative (r = −.11, p.adj < .001) and postoperative EMA compliance (r = −.17, p < .001). Demographic characteristics were robustly associated with EMA compliance, such that individuals who identified as Black/African heritage, used governmentally subsidized insurance, and had less education exhibited lower pre- and postoperative EMA compliance (Table S3-S4). Postoperative EMA compliance was higher among older participants (r=.11, p.adj < .001). In multivariate analyses, the only significant interaction was between sex and race, with Black/African heritage men (n=95) exhibiting low EMA compliance both preoperatively (Median=47.5%, IQR=4-79%) and postoperatively (Median=33.3%, IQR=2-78%; **Fig S3**).

A common concern with EMA is that frequent monitoring could cause symptom worsening (i.e., *reactivity*). We could not evaluate the causal influence of EMA given that everyone was asked to complete EMA, and exposure varied non-randomly based on timing of enrollment and participant compliance. In exploratory analyses, we observed that most participants did not display symptom worsening across the preoperative period (**Fig S4**). Neither receiving more EMAs nor completing a higher proportion of EMAs was associated with greater symptom worsening. Rather, participants who received and completed more EMAs tended to exhibit mild *improvements* in preoperative anxiety (*r* = −.15 and −.12, *p* < .01). These findings should not be interpreted as evidence that EMA reduces anxiety. However, consistent with prior literature, we did not observe substantial preoperative symptom worsening among individuals who completed EMA.

This study establishes the feasibility of perioperative EMA in a large surgical cohort. Without removing individuals based on low compliance, average preoperative compliance with EMA delivered three times daily was 66% (Median=79%), and average postoperative compliance was 60% (Median=71%) in the first 30 days after surgery. These rates were lower than the estimated 75-80% found in numerous meta-analyses across general and specific populations^8–12^. However, 30-50% of EMA studies do not report compliance, and it is common for participants to be excluded from analysis based on unwillingness to complete EMA or low compliance^11, 13, 14^. Consequently, compliance rates reported in the existing literature may be inflated. If we were to remove participants based on no or low compliance (e.g., <20%), our compliance would be consistent with prior meta-analyses. Additionally, postoperative compliance is likely underestimated, as we did not account for times when participants may have been unable to respond (e.g., due to prolonged postoperative sedation). Variation in acute postoperative impairment may account for observed differences in postoperative compliance across surgeries.

Findings may be specific to our study procedures, though there is limited evidence that study-related factors impact compliance^8, 10–12, 15^. Although our sample was larger and more diverse than prior EMA studies^8–12^, there was a lack of representation of races other than White/Caucasian or Black/African heritage. Additionally, because EMA exposure was not randomized, we cannot draw causal conclusions regarding the impact of EMA on subjective symptoms. Future studies should use randomized designs to fully assess potential negative impacts of EMA.

This study is the first to our knowledge to comprehensively assess feasibility of EMA in the perioperative setting. Participants were not paid directly for EMA or excluded from analyses based on low or no EMA completion. Thus, we view our compliance estimates as highly likely to translate to real-world applications of EMA outside of the research context. Overall, remote perioperative monitoring via smartphone appears highly feasible. Integration of EMA into the surgical setting will facilitate novel opportunities to understand perioperative symptom dynamics and monitor risk and recovery factors remotely. Importantly, demographic characteristics including race, insurance status, and education were robustly associated with perioperative EMA compliance. This phenomenon has not previously been investigated in any largescale studies or meta-analyses. Appropriate handling of missing data is critical to ensure models are generalizable to individuals who hold marginalized identities.

## Data Availability

All data produced in the present study are available upon reasonable request to the authors.

## Author Contributions

MRF: Study design; data analysis, manuscript writing and editing; GRM: Data preparation, manuscript writing and editing; KJH: Data preparation, manuscript writing and editing; ZX: Data preparation, manuscript writing and editing; TLR: Manuscript editing; HB: Data collection; KF: Data collection; SA: Data collection; JH: Manuscript editing; CL: Manuscript editing; SH: Study design; funding acquisition; manuscript editing.

