## Supplementary materials for "Remote Perioperative Symptom Monitoring via Smartphone is Feasible: Evidence from the Personalized Prediction of Persistent Postsurgical Pain (P5) study of 2,500 surgical patients"

**Data Collection and Measures**. Baseline data were collected prior to surgery and included: demographics; self-reported lifetime history of anxiety, depression, posttraumatic stress, substance use, and chronic pain conditions; current use of opioid medications; Patient-Reported Outcomes Measurement Information System (PROMIS®) short-form surveys for anxiety, depression, physical functioning, sleep disturbance, and cognitive abilities[15]; Pain Catastrophizing Scale (PCS)[16]; Color-Word Stroop Test (CWST)[17]; and numeric ratings of pain at rest and with movement in the past week on a scale from 0 (no pain) to 10 (worst possible pain). Insurance status and postoperative pain intensity were extracted from the electronic health record. Postoperative pain intensity was defined as the median of all pain ratings collected in the Post Anesthesia Care Unit (PACU), typically within the first 3 hours after surgery.

EMA data were collected using the LifeData app downloaded onto the participant's personal smartphone. The LifeData app is compatible with iOS and Android devices. Participants received EMA prompts three times daily, 5 hours apart. Participants chose to begin receiving EMA prompts at 7am, 8am, or 9am. Each EMA contained 15 items assessing pain intensity, interference, momentary catastrophizing, fatigue, depressed mood, anxiety, and pain medication craving and use. An additional item assessing subjective sleep quality was administered each morning (**Table S1**). Prompts expired 1 hour after delivery. EMA typically began on the day of enrollment and continued through 30 days postoperatively. Participants could self-withdraw from EMA before day 30 by deleting the smartphone application. Participants were not withdrawn from the overall study if they stopped receiving and/or completing EMAs, and compliance was not actively monitored by the study team.

**Statistical Analysis**. Statistical analyses were conducted in R version 4.4. First, we cleaned EMA data collected via LifeData to ensure correct delineation of response vs. non-response. Notably, LifeData appears to categorize EMA as 'responded' if the participant opens the prompt and presses 'submit,' even if no items are completed. Thus, independent data cleaning and verification procedures are necessary. In the current study, an EMA was marked as a response if at least 1 item was completed (i.e., partial completion) within 65 minutes of prompt delivery. This window was chosen because participants were instructed to respond within 1 hour, and it is expected that the participants will take <5 minutes to respond. We removed 1057 observations (.4% of total) that were recorded outside this window. We report average response time across remaining completed prompts. Of the remaining 154,188 observations, 3,119 were partial responses (2% of total). Most partial responses included a response to most items (Median number of missing items = 3). Given the low rate of partial response and relatively low amount of missingness within many partially completed responses, we opted to define compliance with EMA as response to at least 1 item (i.e., partial or full completion).

Next, we compared demographic and clinical characteristics of individuals who did versus did not provide any amount of EMA (1+ survey) using chi-square tests for categorical predictors and nonparametric Wilcoxon rank sum for continuous predictors. We then characterized pre- and postoperative EMA compliance among participants who received any EMA prompts (including individuals who did not complete any prompts). Compliance was calculated for each individual as the number of completed EMAs divided by the total number of possible EMAs in the pre- and postoperative periods. Preoperatively, the total number of possible EMAs was defined as the number of days enrolled prior to surgery times three (for three daily observations). Because some participants had longer preoperative windows due to extenuating circumstances (e.g., delayed surgery), we limited the preoperative window to 14 days before surgery. Postoperatively, all participants had 87 possible EMAs (29 days of three daily observations). A completed EMA was defined as responding to at least 1 EMA item.

For pre- and postoperative compliance, we report the mean, median, interquartile range (IQR), and range across the sample. We also examined demographic and clinical correlates of compliance using nonparametric Wilcoxon rank sum for two-level categorical predictors, Kruskal-Wallis H test for categorical predictors with more than two levels, and Spearman correlations for continuous predictors. Bonferroni correction was used in all analyses to reduce risk of false positives. Effect sizes are reported as $r$ for Wilcoxon rank sum and Spearman correlations, where 0.1, 0.3, and 0.5 indicate small, medium, and large effects, respectively. Effect sizes are reported as $\eta^2$ for Kruskal-Wallis, where 0.01, 0.06, and 0.14 indicate small, medium, and large effects, respectively[18].

We then used random forest models to explore possible interactions among predictors of compliance. Given small cell sizes in some categories, only participants who identified their race as Black/African heritage or White/Caucasian were included. Missing predictor values were imputed using the missForest algorithm. The imputed dataset was used to train a random forest regression with default parameters (500 trees, $\sqrt{p}$ variables considered at each split). Model performance was estimated using out-of-bag (OOB) error and pseudo $R^2$. To explore potential interactions among predictors, pairwise interaction strengths were extracted using the find.interaction() function.

A common concern with EMA is that frequent monitoring could cause symptom worsening (i.e., *reactivity*). We could not evaluate the causal influence of EMA given that all participants received EMA. However, there was variation in how many EMAs participants received and completed, based on timing of enrollment and compliance. In an exploratory analysis, we examined these metrics of EMA exposure as correlates of preoperative, within-person change across symptom domains. First, we used linear mixed-effects models to extract a person-specific random slope estimate for each EMA symptom domain (pain, depressed mood, anxiety, catastrophizing). We then examined Pearson correlations between these estimates and metrics of EMA exposure (number of EMA received; number of EMA completed).

**Table S1. Ecological Momentary Assessment (EMA) Items**

1. To what extent did you have trouble sleeping last night?*
2. Right now, how intense is your pain?
3. Right now, how much is pain interfering with your enjoyment of life?
4. Right now, how much is pain interfering with your activities?
5. Right now, I keep thinking about how much I hurt.
6. Right now, my pain overwhelms me.
7. Right now, I am afraid that my pain will get worse.
8. Right now, how tired are you feeling?
9. Right now, how depressed are you feeling?
10. Right now, how hopeless are you feeling?
11. Right now, how worthless are you feeling?
12. Right now, how tense are you feeling?
13. Right now, how anxious are you feeling?
14. Right now, how nervous are you feeling?
15. Right now, how badly do you feel you need pain medications?
16. Since your last prompt, how much pain medication have you used?

*Note.* All items were presented in randomized order at each prompt, except for item 1 (trouble sleeping), which was only asked on the first survey of the day. All items were responded to on a scale from 0 ("not at all") to 100 ("worst possible"). The average EMA response time was 89 seconds (Median = 65 seconds, SD = 137 seconds).

**Table S2. Demographic and clinical characteristics of participants who did and did not complete any EMA**

| Predictor | Levels | Has EMA<br>*N = 2100 (90%)* | No EMA<br>*N = 229 (10%)* | p | p.adj |
|---|---|---|---|---|---|
| **Race** | American Indian/Alaskan Native | 15 (1) | 1 (0) | 0.204 | 1 |
| | Asian | 14 (1) | | | |
| | Black/African Heritage | 372 (18) | 55 (24) | | |
| | White/Caucasian | 1594 (76) | 158 (70) | | |
| | Hawaiian Native/Other Pacific Islander | 4 (0) | | | |
| | More than one | 33 (2) | 5 (2) | | |
| | Other | 30 (1) | 2 (1) | | |
| | Prefer not to answer | 23 (1) | 4 (2) | | |
| **Ethnicity** | Hispanic | 47 (2) | 4 (2) | 0.851 | 1 |
| | Non-Hispanic | 1961 (98) | 209 (98) | | |
| **Age** | Median (IQR) | 54.25 (42.73 to 64.39) | 58.62 (45.83 to 67.49) | 0.0005 | **0.011** |
| **Sex** | Female | 1419 (68) | 136 (59) | 0.015 | 0.339 |
| | Male | 681 (32) | 93 (41) | | |
| **Education** | College degree | 450 (22) | 40 (18) | 0.079 | 1 |
| | High school diploma | 470 (22) | 62 (28) | | |
| | Less than high school | 112 (5) | 18 (8) | | |
| | Masters degree | 297 (14) | 21 (9) | | |
| | Post-graduate degree | 89 (4) | 11 (5) | | |
| | Some college | 673 (32) | 72 (32) | | |
| **Marital Status** | Divorced | 236 (11) | 34 (15) | 0.063 | 1 |
| | Domestic partnership | 41 (2) | 10 (4) | | |
| | Married | 1142 (55) | 107 (48) | | |
| | Other | 8 (0) | | | |
| | Single | 505 (24) | 58 (26) | | |
| | Widowed | 106 (5) | 12 (5) | | |
| | Separated | 46 (2) | 3 (1) | | |
| **Anxiety** | Yes | 859 (41) | 95 (42) | 0.837 | 1 |
| **Depression** | Yes | 786 (38) | 98 (44) | 0.095 | 1 |
| **PTSD** | Yes | 226 (11) | 26 (12) | 0.822 | 1 |
| **SUD** | Yes | 68 (3) | 12 (5) | 0.159 | 1 |
| **Chronic Pain** | Yes | 630 (30) | 75 (34) | 0.35 | 1 |
| **Current Opioid Use** | Yes | 276 (14) | 41 (19) | 0.046 | 1 |
| **Pain at Rest** | Median (IQR) | 3.00 (1.00 to 6.00) | 4.00 (1.00 to 7.00) | 0.004 | 0.078 |
| **Pain with Activity** | Median (IQR) | 5.00 (2.00 to 7.00) | 5.00 (2.00 to 8.00) | 0.012 | 0.272 |
| **PCS** | Median (IQR) | 8.00 (2.00 to 18.00) | 10.00 (4.00 to 21.00) | 0.02 | 0.429 |
| **PROMIS Cognition** | Median (IQR) | 49.60 (43.80 to 58.80) | 50.75 (42.60 to 57.40) | 0.335 | 1 |
| **PROMIS PF** | Median (IQR) | 41.00 (34.70 to 48.60) | 38.65 (34.18 to 45.70) | 0.019 | 0.424 |
| **PROMIS Anxiety** | Median (IQR) | 52.50 (40.30 to 59.40) | 54.10 (40.30 to 60.30) | 0.198 | 1 |
| **PROMIS Depression** | Median (IQR) | 48.90 (41.00 to 55.90) | 49.00 (41.00 to 57.20) | 0.042 | 0.925 |
| **PROMIS Sleep** | Median (IQR) | 52.80 (46.40 to 58.10) | 52.70 (48.18 to 56.73) | 0.862 | 1 |
| **Stroop** | Median (IQR) | 50.00 (45.00 to 54.00) | 47.00 (44.00 to 52.00) | 0.006 | 0.132 |
| **Surgical Site** | Breast | 57 (3) | 4 (2) | 0.053 | 1 |
| | Cardiothoracic | 213 (10) | 32 (14) | | |
| | GI | 445 (21) | 45 (20) | | |
| | GUR | 618 (29) | 55 (24) | | |
| | Head & Neck | 121 (6) | 8 (3) | | |

| | | | | | |
|---|---|---|---|---|---|
| | Ortho | 372 (18) | 49 (21) | | |
| | Other | 19 (1) | 6 (3) | | |
| | Spine | 226 (11) | 26 (11) | | |
| | Vascular | 29 (1) | 4 (2) | | |
| **Insurance** | Managed Care | 975 (47) | 69 (30) | <0.00001 | **<.0001** |
| | Medicaid | 227 (11) | 42 (19) | | |
| | Medicare | 585 (28) | 89 (39) | | |
| | Other | 95 (5) | 6 (3) | | |
| | Self-Pay | 212 (10) | 21 (9) | | |

*Note*. p.adj = Bonferroni-corrected p-value; PTSD = Post-Traumatic Stress Disorder; SUD = Substance Use Disorder; PCS = Pain Catastrophizing Scale; PROMIS = Patient-Reported Outcomes Measurement Information System; PF = Physical Function; Stroop = Color-Word Stroop Test. Anxiety, depression, PTSD, SUD, and chronic pain refer to self-reported lifetime diagnoses. Past week pain with activity and pain at rest were assessed via 0-10 numeric rating scales.

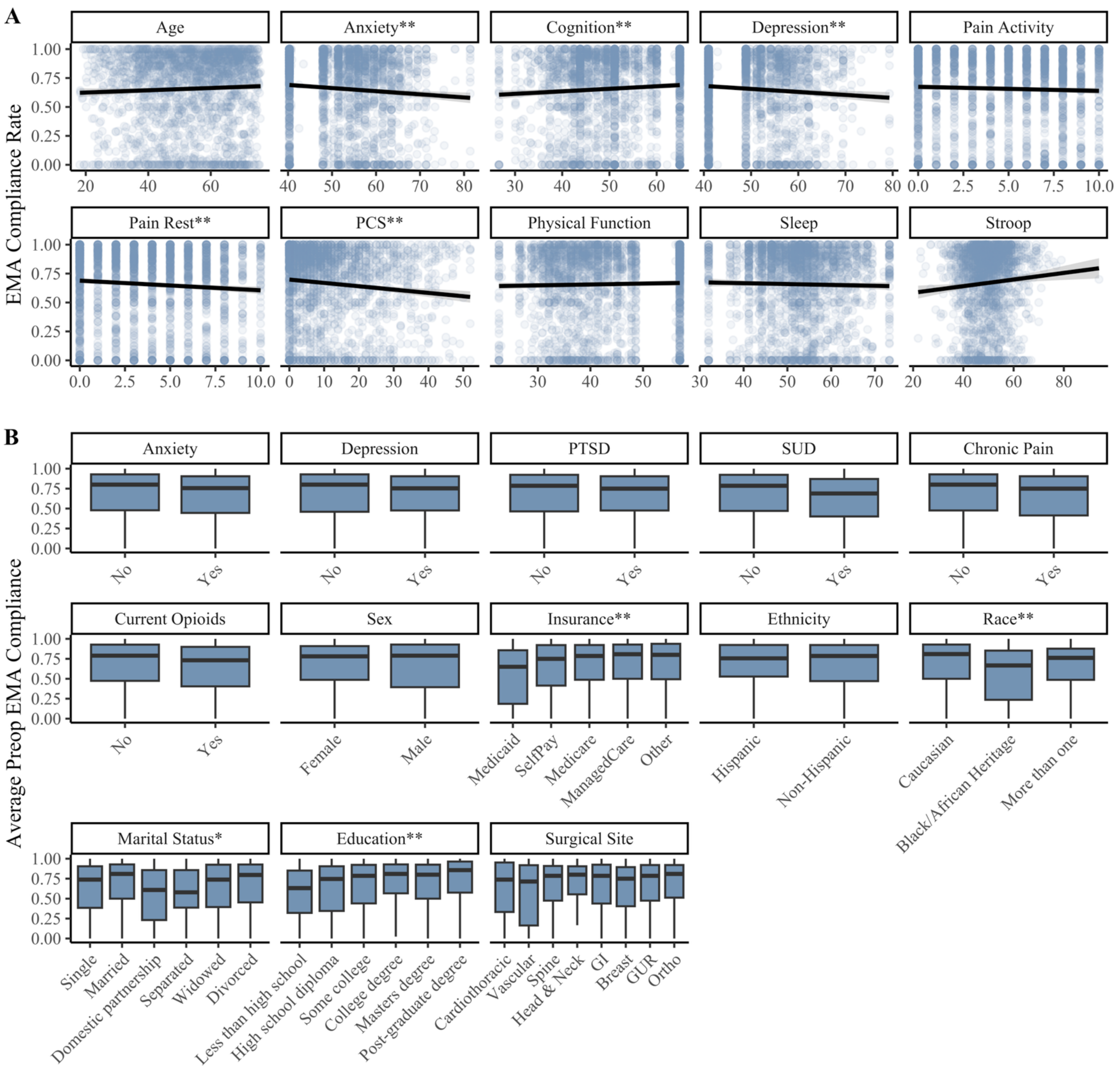


**Figure S1.** Associations between preoperative Ecological Momentary Assessment (EMA) compliance and demographic and clinical characteristics. Panel **A** shows correlations between preoperative compliance and continuous predictors. Panel **B** shows box plots of preoperative compliance by categorical predictors (midline = median). **Bonferroni-corrected p < .01; *Bonferroni-corrected p < .05; PCS = Pain Catastrophizing Scale; PTSD = Post-Traumatic Stress Disorder; SUD = Substance Use Disorder; GI = gastrointestinal; GUR = Genitourinary. In Panel A, anxiety, subjective cognition, depression, physical function, and subjective sleep impairment were assessed via PROMIS self-report measures; pain with activity and pain at rest were assessed via numeric rating scales. In Panel B, anxiety, depression, PTSD, SUD, and chronic pain refer to self-reported lifetime diagnoses.

**Table S3. Predictors of preoperative EMA compliance**

| Predictor | Levels | n (%) | Median | Mean | IQR | $\eta^2$ | p.adj |
|---|---|---|---|---|---|---|---|
| **Anxiety Dx** | No | 1268 (59%) | 80 | 67.1 | 48, 93 | 0.003 | 0.513 |
| | Yes | 891 (41%) | 75.6 | 64 | 45, 90 | | |
| **Depression Dx** | No | 1328 (62%) | 80 | 66.5 | 46, 93 | 0.002 | 1 |
| | Yes | 820 (38%) | 75.3 | 64.4 | 48, 90 | | |
| **PTSD Dx** | No | 1909 (89%) | 78.6 | 65.9 | 46, 92 | | |
| | Yes | 239 (11%) | 75 | 64.1 | 48, 90 | | |
| **SUD Dx** | No | 2070 (97%) | 78.6 | 66 | 47, 92 | | |
| | Yes | 74 (3%) | 68.9 | 59.9 | 40, 87 | | |
| **Chronic Pain** | No | 1496 (70%) | 80 | 66.7 | 48, 93 | 0.003 | 0.787 |
| | Yes | 649 (30%) | 75 | 63.7 | 41, 90 | | |
| **Current Opioid Use** | No | 1815 (86%) | 78.9 | 66.3 | 47, 93 | 0.003 | 0.856 |
| | Yes | 289 (14%) | 73.2 | 62.6 | 40, 90 | | |
| **Sex** | Female | 1456 (67%) | 78.1 | 66.4 | 49, 91 | 0 | 1 |
| | Male | 715 (33%) | 78.9 | 64.3 | 40, 93 | | |
| **Insurance** | Medicaid | 246 (11%) | 65 | 55.5 | 19, 86 | 0.014 | **<.001** |
| | Self-Pay | 219 (10%) | 75 | 65 | 41, 92 | | |
| | Medicare | 615 (28%) | 78.6 | 65.9 | 49, 92 | | |
| | Managed Care | 989 (46%) | 80.8 | 68.1 | 50, 93 | | |
| | Other | 96 (4%) | 80 | 67.2 | 49, 94 | | |
| **Ethnicity** | Hispanic | 50 (2%) | 75.6 | 68.1 | 53, 92 | 0 | 1 |
| | Non-Hispanic | 2019 (98%) | 78.6 | 66 | 47, 92 | | |
| **Race** | White/Caucasian | 1639 (79%) | 81 | 68.4 | 50, 93 | 0.029 | **<.001** |
| | Black/African Heritage | 394 (19%) | 66.7 | 55.6 | 24, 85 | | |
| | More than one | 35 (2%) | 76.2 | 63.7 | 49, 88 | | |
| **Marital Status** | Single | 528 (25%) | 73.8 | 62 | 38, 90 | 0.011 | **0.022** |
| | Married | 1166 (54%) | 81 | 68.4 | 50, 93 | | |
| | Domestic partnership | 47 (2%) | 60.9 | 55.6 | 23, 86 | | |
| | Separated | 45 (2%) | 57.9 | 57.1 | 39, 86 | | |
| | Widowed | 109 (5%) | 73.8 | 62.8 | 39, 92 | | |
| | Divorced | 248 (12%) | 79.7 | 66 | 45, 93 | | |
| **Education** | Less than high school | 116 (5%) | 63.2 | 55.4 | 32, 85 | 0.017 | **<.001** |
| | High school diploma | 490 (23%) | 74.6 | 61.9 | 35, 90 | | |
| | Some college | 699 (32%) | 78.6 | 65 | 44, 92 | | |
| | College degree | 459 (21%) | 81 | 70.8 | 57, 93 | | |
| | Master's degree | 304 (14%) | 80 | 68.6 | 50, 93 | | |
| | Post-graduate degree | 90 (4%) | 85.7 | 71.2 | 58, 96 | | |
| **Surgical Site** | Cardiothoracic | 232 (11%) | 73.8 | 62.8 | 33, 95 | 0.007 | 1 |
| | Vascular | 31 (1%) | 71.4 | 59 | 16, 92 | | |
| | Spine | 231 (11%) | 78.6 | 65.9 | 47, 91 | | |
| | Head & Neck | 122 (6%) | 80 | 70.8 | 55, 90 | | |
| | Gastrointestinal | 456 (21%) | 78.6 | 65.4 | 44, 92 | | |
| | Breast | 57 (3%) | 75 | 60.8 | 40, 89 | | |
| | Genitourinary | 632 (29%) | 78.6 | 66.4 | 47, 92 | | |
| | Orthopedic | 385 (18%) | 81 | 67.6 | 51, 92 | | |

*Note*. IQR = interquartile range; p.adj = Bonferroni-corrected p-value; Dx = self-reported lifetime diagnoses; PTSD = Post-Traumatic Stress Disorder; SUD = Substance Use Disorder.

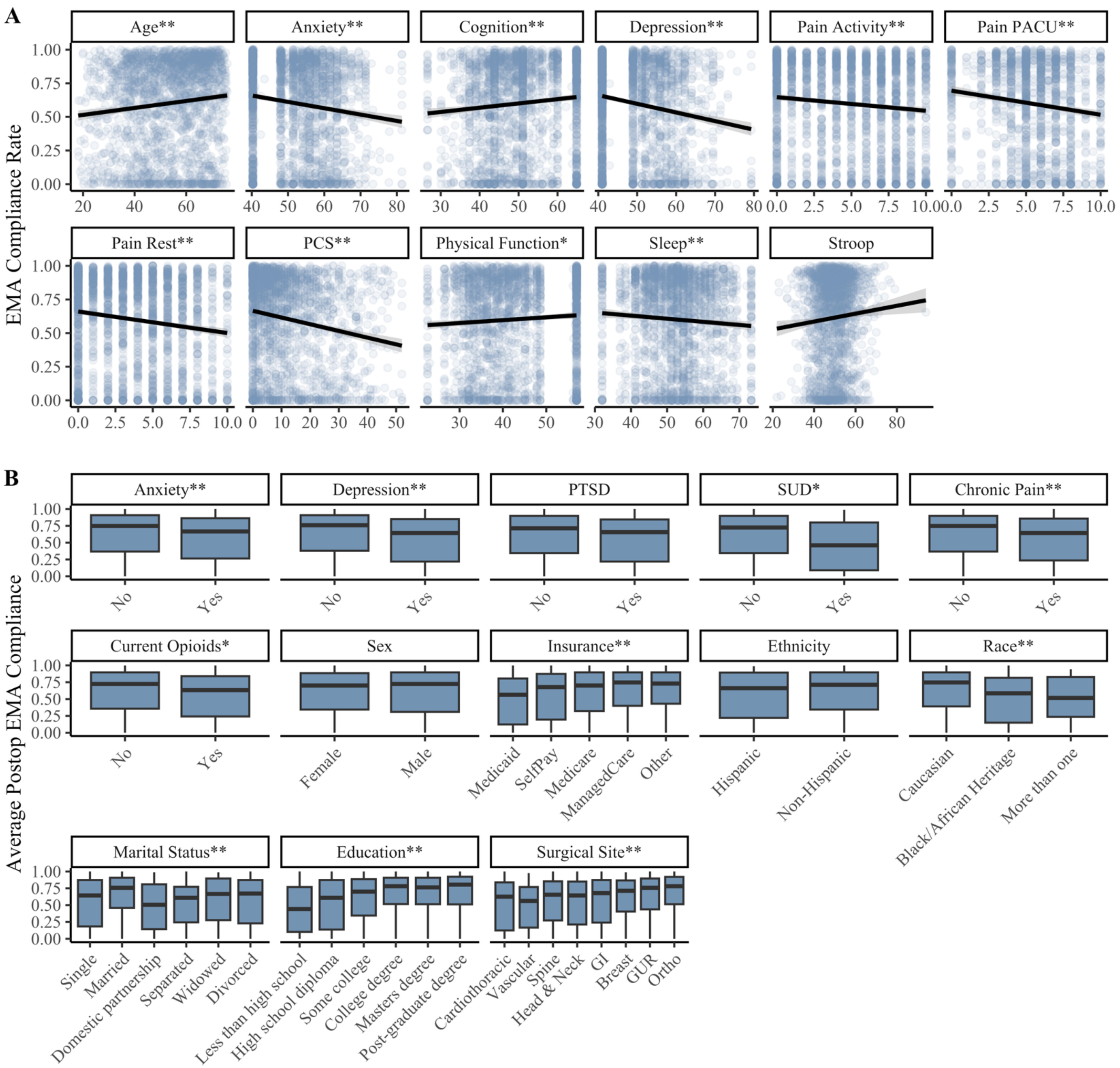


**Figure S2.** Associations between postoperative Ecological Momentary Assessment (EMA) compliance and demographic and clinical characteristics. Panel **A** shows correlations between postoperative compliance and continuous predictors. Panel **B** shows box plots of postoperative compliance by categorical predictors (midline = median). **Bonferroni-corrected $p < .01$; *Bonferroni-corrected $p < .05$; PACU = Post Anesthesia Care Unit; PCS = Pain Catastrophizing Scale; PTSD = Post-Traumatic Stress Disorder; SUD = Substance Use Disorder; GI = gastrointestinal; GUR = Genitourinary. In Panel A, anxiety, subjective cognition, depression, physical function, and subjective sleep impairment were assessed via PROMIS self-report measures; pain with activity and pain at rest were assessed via numeric rating scales. In Panel B, anxiety, depression, PTSD, SUD, and chronic pain refer to self-reported lifetime diagnoses.

**Table S4. Predictors of postoperative EMA compliance**

| Predictor | Levels | n (%) | Median | Mean | IQR | $\eta^2$ | p.adj |
|---|---|---|---|---|---|---|---|
| **Anxiety Dx** | No | 1249 (59%) | 74.7 | 62.3 | 37, 91 | 0.009 | **0.001** |
| | Yes | 869 (41%) | 66.7 | 57 | 26, 86 | | |
| **Depression Dx** | No | 1303 (62%) | 75.9 | 62.9 | 38, 91 | 0.014 | **<.001** |
| | Yes | 804 (38%) | 64.4 | 55.7 | 22, 85 | | |
| **PTSD Dx** | No | 1873 (89%) | 71.3 | 60.6 | 34, 90 | 0.004 | 0.304 |
| | Yes | 235 (11%) | 65.5 | 55.3 | 22, 84 | | |
| **SUD Dx** | No | 2031 (97%) | 72.4 | 60.6 | 34, 90 | 0.005 | **0.039** |
| | Yes | 72 (3%) | 46 | 45.8 | 9, 80 | | |
| **Chronic Pain** | No | 1473 (70%) | 74.7 | 61.9 | 37, 90 | 0.009 | **0.001** |
| | Yes | 631 (30%) | 64.4 | 55.8 | 24, 86 | | |
| **Current Opioid Use** | No | 1784 (86%) | 72.4 | 61.2 | 36, 90 | 0.006 | **0.02** |
| | Yes | 280 (14%) | 63.2 | 54.8 | 24, 84 | | |
| **Sex** | Female | 1429 (67%) | 70.1 | 60.2 | 34, 89 | 0 | 1 |
| | Male | 701 (33%) | 72.4 | 59.7 | 31, 90 | | |
| **Insurance** | Medicaid | 230 (11%) | 56.3 | 49.5 | 12, 80 | 0.015 | **<.001** |
| | Self-Pay | 217 (10%) | 67.8 | 56.7 | 20, 87 | | |
| | Medicare | 599 (28%) | 70.1 | 59.9 | 32, 90 | | |
| | Managed Care | 984 (46%) | 74.7 | 63 | 40, 90 | | |
| | Other | 94 (4%) | 73 | 62.6 | 43, 90 | | |
| **Ethnicity** | Hispanic | 50 (2%) | 66.1 | 55.6 | 22, 89 | 0 | 1 |
| | Non-Hispanic | 1982 (98%) | 71.3 | 60.6 | 34, 90 | | |
| **Race** | White/Caucasian | 1613 (80%) | 74.7 | 62.6 | 39, 90 | 0.022 | **<.001** |
| | Black/African Heritage | 379 (19%) | 58.6 | 50.4 | 15, 82 | | |
| | More than one | 34 (2%) | 51.7 | 50 | 24, 83 | | |
| **Marital Status** | Single | 511 (24%) | 64.4 | 54.2 | 18, 87 | 0.020 | **<.001** |
| | Married | 1153 (55%) | 75.9 | 64.6 | 46, 91 | | |
| | Domestic partnership | 43 (2%) | 50.6 | 49 | 14, 81 | | |
| | Separated | 46 (2%) | 60.9 | 53.4 | 24, 77 | | |
| | Widowed | 109 (5%) | 66.7 | 56.6 | 28, 90 | | |
| | Divorced | 240 (11%) | 67.2 | 56.8 | 23, 87 | | |
| **Education** | Less than high school | 114 (5%) | 44.3 | 44.7 | 10, 77 | 0.034 | **<.001** |
| | High school diploma | 472 (22%) | 60.9 | 52.1 | 14, 87 | | |
| | Some college | 682 (32%) | 70.1 | 59.8 | 34, 89 | | |
| | College degree | 454 (21%) | 78.2 | 66.6 | 52, 91 | | |
| | Master's degree | 304 (14%) | 76.4 | 66.1 | 51, 91 | | |
| | Post-graduate degree | 92 (4%) | 80.5 | 69.5 | 51, 92 | | |
| **Surgical Site** | Cardiothoracic | 224 (11%) | 62.6 | 51.3 | 12, 84 | 0.029 | **<.001** |
| | Vascular | 31 (1%) | 56.3 | 49.4 | 17, 77 | | |
| | Spine | 228 (11%) | 65.5 | 56.8 | 27, 85 | | |
| | Head & Neck | 122 (6%) | 64.4 | 55.5 | 21, 85 | | |
| | Gastrointestinal | 445 (21%) | 67.8 | 57.4 | 24, 87 | | |
| | Breast | 58 (3%) | 71.3 | 61.3 | 41, 87 | | |
| | Genitourinary | 617 (29%) | 75.9 | 64.1 | 44, 90 | | |
| | Orthopedic | 380 (18%) | 78.2 | 66.9 | 51, 92 | | |

*Note*. IQR = interquartile range; p.adj = Bonferroni-corrected p-value; Dx = self-reported lifetime diagnoses; PTSD = Post-Traumatic Stress Disorder; SUD = Substance Use Disorder.

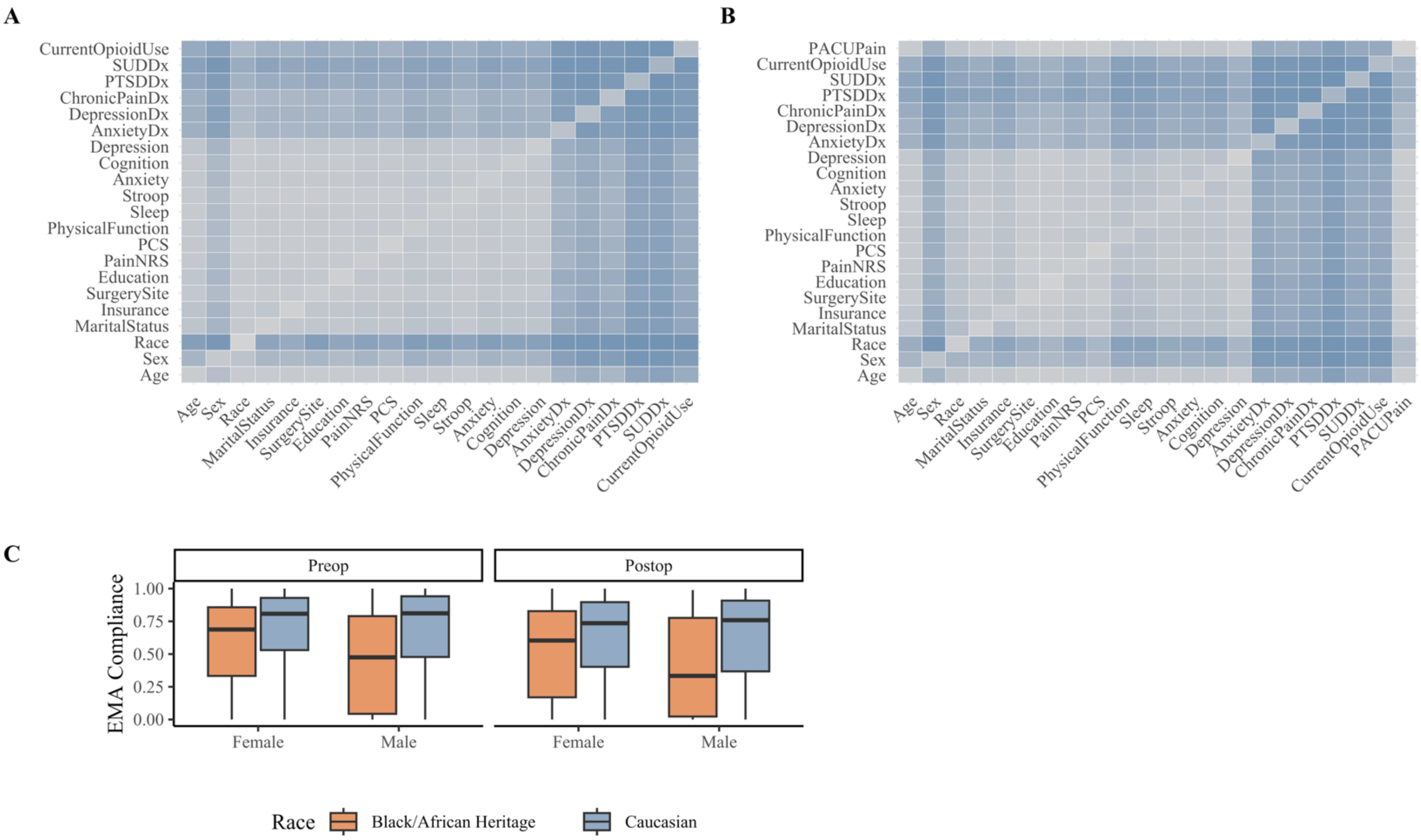


**Figure S3. Interaction effects among predictors of Ecological Momentary Assessment (EMA) compliance**. Panel **A** is a heatmap of interaction strength derived from a random forest regression model predicting preoperative EMA compliance (grey = low strength; blue = higher strength). Panel **B** is a heatmap of interaction strength derived from a random forest regression model predicting postoperative EMA compliance. Panel **C** displays the only statistically significant interaction (race x sex) in follow-up linear regression analysis. PACU = Post Anesthesia Care Unit; PCS = Pain Catastrophizing Scale; PTSD = Post-Traumatic Stress Disorder; SUD = Substance Use Disorder; NRS = numeric rating scale (average of pain at rest and pain with activity). Predictors labeled 'Dx' refer to lifetime diagnoses. Predictors labeled 'Depression,' 'Cognition,' 'Anxiety,' 'Sleep,' and 'PhysicalFunction' are PROMIS measures.

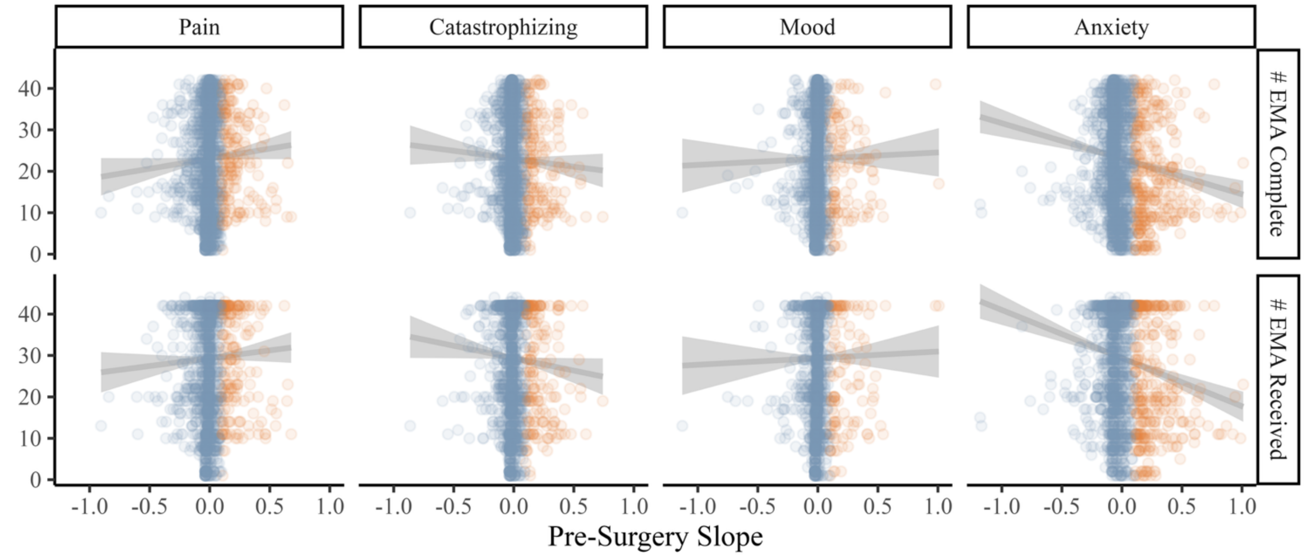


**Fig S4. Exploratory analysis of reactivity to EMA**. Top row shows associations between the number of EMAs completed and pre-surgery symptom severity slope (orange = random slope > .1, suggesting symptom worsening). Bottom row shows association between the number of EMAs received and pre-surgery symptom severity slope. Results suggest neither receiving nor completing more EMAs was associated with symptom worsening. Rather, individuals who received and completed more EMAs tended to exhibit mild improvements in anxiety symptoms ($r$ = -.15 and -.12, $p < .01$). Constructs are composites of relevant EMA items (Pain = items 2-4; Catastrophizing = items 4-7; Mood = items 8-11; Anxiety = items 12-14 in Table S1).
